# Distinct impact modes of polygenic disposition to dyslexia in the adult brain

**DOI:** 10.1101/2024.02.14.24302815

**Authors:** Sourena Soheili-Nezhad, Dick Schijven, Rogier B. Mars, Simon E. Fisher, Clyde Francks

**Affiliations:** Language and Genetics Department, Max Planck Institute for Psycholinguistics, Nijmegen, The Netherlands; Donders Institute for Brain, Cognition and Behaviour, Radboud University, Nijmegen, the Netherlands; Wellcome Centre for Integrative Neuroimaging, Centre for Functional MRI of the Brain (FMRIB), Nuffield Department of Clinical Neurosciences, John Radcliffe Hospital, University of Oxford, Oxford, UK; Department of Cognitive Neuroscience, Radboud University Medical Center, Nijmegen, the Netherlands

**Keywords:** Dyslexia, polygenic score, general population, tensor-based morphometry, fixel-based analysis

## Abstract

Dyslexia is a common condition that impacts reading ability. Identifying affected brain networks has been hampered by limited sample sizes of imaging case-control studies. We focused instead on brain structural correlates of genetic disposition to dyslexia in large-scale population data. In over 30,000 adults (UK Biobank), higher polygenic disposition to dyslexia was associated with lower head and brain size, and especially reduced volume and/or altered fiber density in networks involved in motor control, language and vision. However, individual genetic variants disposing to dyslexia often had quite distinct patterns of association with brain structural features. Independent component analysis applied to brain-wide association maps for thousands of dyslexia-disposing genetic variants revealed multiple impact modes on the brain, that corresponded to anatomically distinct areas with their own genomic profiles of association. Polygenic scores for dyslexia-related cognitive and educational measures, as well as attention-deficit/hyperactivity disorder, showed similarities to dyslexia polygenic disposition in terms of brain-wide associations, with microstructure of the internal capsule consistently implicated. In contrast, lower volume of the primary motor cortex was only associated with higher dyslexia polygenic disposition among all traits. These findings robustly reveal heterogeneous neurobiological aspects of dyslexia genetic disposition, and whether they are shared or unique with respect to other genetically correlated traits.

## Introduction

Roughly 3-7% of school-age children have dyslexia, a neurodevelopmental condition that affects reading, writing and spelling^1^. Reading acquisition during childhood is accompanied by the adaptation of several brain networks, and multiple hypotheses have been formulated to explain the etiology of dyslexia through altered developmental trajectories in these networks and the functions they support^2^. The phonological deficit hypothesis suggests that dyslexia involves diminished ability in associating *phonemes*—the units of spoken language—with written linguistic symbols or *graphemes*, sometimes stemming from a lack of awareness of the phonological structure of language^3^. In contrast, the orthographic deficit hypothesis suggests that some dyslexic readers may not identify words as cohesive patterns, but instead decode them as sequences of letters at a slow pace due to impairments of the visual stream^4^. Yet other mechanistic models highlight auditory^5^ and magnocellular^6^ pathways. Impairments of rapid automatized naming^7^, verbal short-term memory^8^ and attention control have also been implicated^9, 10^. Rather than there being a single, monolithic explanation for dyslexia, it is likely that the underlying mechanisms are heterogeneous and multifactorial^11, 12^.

Functional neuroimaging of people with dyslexia has suggested reduced activation or functional connectivity during reading-related tasks of various left-hemisphere regions that are important for language and/or normal reading, including the posterior temporo-parietal cortex, the inferior frontal gyrus, and the anterior occipito-temporal cortex^13–15^ ^16, 17^. However, these efforts often employed divergent methods and task paradigms in sample sizes of only tens of individuals, and findings have often been inconsistent^18, 19^. In terms of brain structural MRI too, results from multi-cohort or meta-analysis studies in total sample sizes up to hundreds of individuals have not aligned well, yielding negative findings or much smaller effects than originally reported in smaller individual studies^20–23^. Moreover, the largest diffusion MRI-based investigation of white-matter microstructure, in 104 affected children and adolescents compared to 582 controls, did not detect significant group-wise differences^24^.

This overall sequence has been encountered in neuroimaging studies of multiple other traits beyond dyslexia; initial waves of underpowered, hypothesis-driven studies produced inconsistent results, followed by larger more systematic screening studies that failed to replicate the initial findings, while sometimes producing unanticipated new leads^25^. Taken together, it is clear that hypothesis-free brain-wide mapping in much larger sample sizes is needed, to better understand the brain regions and networks involved in dyslexia^26^.

The heritability of dyslexia is estimated to be roughly 40-70% based on twin studies^27, 28^, with common DNA variants accounting for around 15% of its disposition according to genome-wide investigations^29,30^. Dyslexia also shows substantial genetic correlations (in the range of 0.6 to 0.8) with measures of reading and spelling performance, and phonemic awareness, more broadly across the population^31^. Here, we reasoned that estimating polygenic disposition to dyslexia in the UK Biobank, a large general population dataset where genome-wide genotype and neuroimaging data are available^32–34^, would reveal neurobiological markers relevant to the development and/or manifestation of dyslexia. To calculate polygenic disposition in the UK Biobank individuals, we made use of genome-wide association summary statistics from a recent study of 51,800 individuals who reported having received a dyslexia diagnosis, and over one million controls, carried out by 23andMe, Inc.^30^.

We carried out our brain mapping analyses with respect to voxel-wise volumetric measures, as well as fixel-wise apparent fiber density, the latter for the analysis of white matter microstructure. Different predisposing genetic loci may impact distinct brain regions and networks. We therefore aimed to disentangle heterogeneity in the brain-wide associations of different dyslexia disposing variants, by decomposing the overall polygenic disposition into a number of distinct impact modes in terms of neurobiological correlates. For this, we developed a novel application of independent component analysis^35, 36^.

Dyslexia is also associated with several other traits related to cognition, education and behaviour, and shows significant genetic correlations with attention-deficit hyperactivity disorder (ADHD), educational attainment, and intelligence^30^. This means that some of the genetic factors that predispose to dyslexia are shared with these other traits. The question then arises: which structural brain features are associated with polygenic disposition to dyslexia alone, versus more generally with polygenic dispositions to a range of cognitive, educational and behavioural traits that are associated with dyslexia? The combination of brain features uniquely associated with dyslexia polygenic disposition is likely to distinguish liability to this particular trait among others. We therefore went on to quantify the polygenic dispositions of UK Biobank individuals to ADHD, educational attainment, school grades, fluid intelligence, and the reading-related psychometric traits of single-word reading, non-word reading, spelling, and phonemic awareness^31^. We mapped the brain structural correlates of all of these polygenic dispositions in the UK Biobank, and compared and contrasted with the brain maps for dyslexia polygenic disposition.

## Results

### Brain correlates of dyslexia polygenic scores

After genetic and brain imaging quality control, we generated dyslexia PGS for between 31,695 and 35,231 adult individuals from the UK Biobank dataset, depending on the availability of data for diffusion and T1-weighted MRI modalities, respectively (see Methods). We mapped brain wide associations of dyslexia PGS with voxel-wise regional volume derived from tensor-based morphometry^37, 38^, as well as microstructural measure of apparent fiber density derived from fixel-based analysis^39^ (Methods). For our main analysis, we report results for PGS generated with lassosum2^40^ that were optimized for capturing inter-individual brain variation (Methods; Supplementary Fig. 1), but other automated polygenic methods including SBayesR^41^ and PRS-CS^42^ delivered highly comparable results (Supplementary Fig. 2).

Individuals with higher dyslexia PGS exhibited lower total brain volume, which was more apparent in gray matter than white matter (t=-6.6 and t=-5.5, respectively; Supplementary Table 1). Among other global measures of brain anatomy, dyslexia PGS was most strongly associated with lower total cortical surface area, especially of the left hemisphere (t=-6.4). Unexpectedly, dyslexia PGS was slightly more predictive of overall head size (t=-6.9), which is a measure derived from skull anatomy^43^, than any brain measure (Supplementary Table 1). Head size or total brain volume are commonly used as unwanted covariates in structural neuroimaging analysis. However, we reasoned that adjusting for head size could introduce collider bias in voxel-based volumetric analysis. Collider bias can occur when adjusting for a supposed confound variable, head size in this case, that is actually influenced by both variables between which an association is tested – here dyslexia PGS as the predictor and brain regional volume as outcome. Therefore, we performed our primary voxel-wise analysis without this adjustment (i.e. using raw Jacobian determinant values comprising both the nonlinear and linear registration components), but also repeated the analysis secondarily with head size adjustment.

Without head size adjustment, individuals with higher dyslexia PGS showed lower regional volumes across multiple brain regions after brain-wide multiple comparison correction (Methods). This included a large frontal cluster along the medial wall, extending from Brodmann area 4 to perigenual medial frontal cortex (Fig. 1: top). Additionally, lower volume was observed in midbrain, thalamus, and bilateral amygdalae, in individuals with higher dyslexia PGS (Fig. 1: top). Many of these associations were evident in both hemispheres and more or less bilaterally symmetric, except two clusters of lateralized lower volume in the left anterior insula and in the left posterior temporoparietal junction, again associated with higher dyslexia PGS (Fig. 1: top).

**Figure 1.**
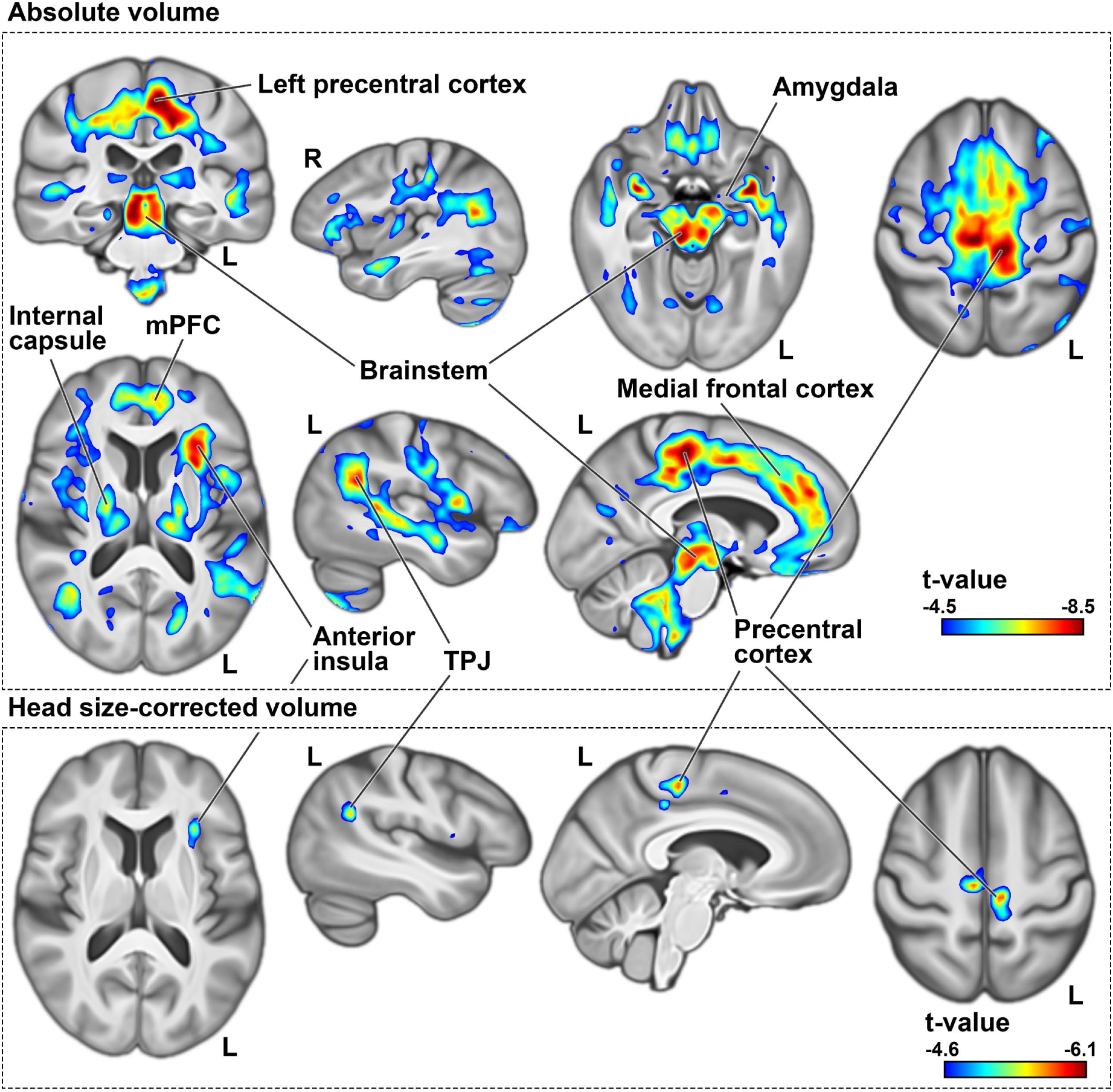
Negative dyslexia PGS associations with regional brain volume before (top) and after head size correction (bottom). Clusters indicate voxels whose volumes are significantly lower in individuals with higher polygenic disposition to dyslexia, at p-values of smaller than 0.05 as obtained from non-parametric testing, with brain-wide correction for multiple comparisons using 5000 permutations. In these significant clusters, voxels are coloured based on t-values derived from similar parametric tests, in order to visualize effect sizes and peak regions. All figures are shown in radiological convention, where the left side in transverse and coronal views corresponds to the right cerebral hemisphere and vice versa. R: right. L: left. TPJ: temporo-parietal junction.

There were no regions where higher dyslexia PGS was associated with higher regional volumes in voxel-wise analysis after multiple comparison correction, although there were sub-threshold (non-significant) positive associations between dyslexia PGS and the volumes of bilateral primary visual cortices and middle temporal gyri (Supplementary Fig. 3).

Following adjustment for head size as a covariate, negative associations were again observed between dyslexia PGS and similar brain regions as the non-adjusted analysis, although the significant clusters were markedly smaller compared to the previous analysis (Fig. 1: bottom). Notably, the sub-threshold (i.e. nonsignificant) positive voxel-wise associations with the primary visual cortex and anterior middle temporal gyrus became significant after head size correction, indicating that individuals with higher dyslexia PGS had higher volumes *relative to their head size* in these regions (Supplementary Fig. 3).

In fixel-wise analysis of white-matter microstructure, dyslexia PGS was positively associated with apparent fiber density in forceps major tracts, which connect homologous regions of the bilateral occipital cortices (Fig. 2: top). In contrast, dyslexia PGS was negatively associated with apparent fiber density in three separate clusters of fixels bilaterally: within the superior longitudinal fasciculi, cerebellar dentate nuclei, and anterior limb of the internal capsule (Fig. 2: top). Fiber tractography revealed that the internal capsule and dentate fixels highlight tracts that pass through the brainstem and superior cerebellar peduncles, with neocortical connections that mainly span the frontal and parietal cortices (Figure 2: bottom). (Note that adjustment for head size is not relevant for fixel-based analysis of white-matter microstructure.)

**Figure 2.**
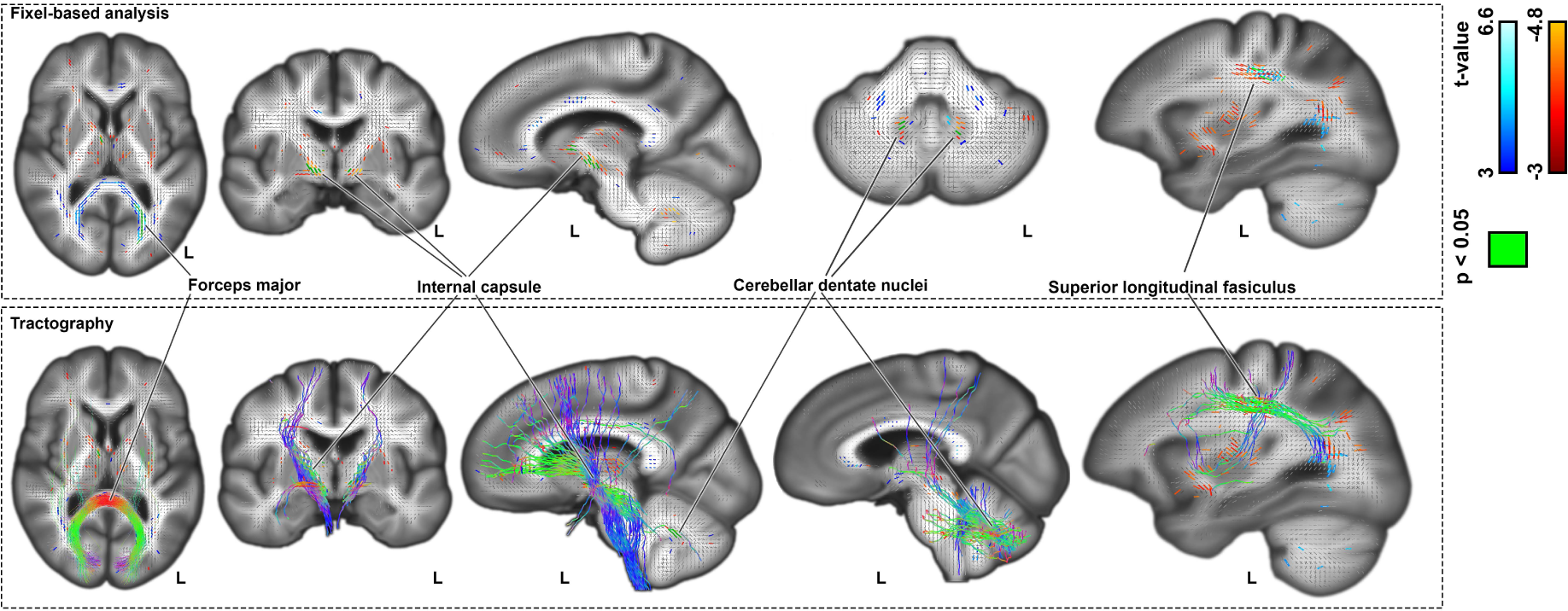
Top: association of dyslexia PGS with fixel-wise apparent fiber density. Bottom: Probabilistic tractography streamlines coloured based on fiber directions. Figures are shown in radiological convention, where the left side in transverse and coronal views corresponds to the right cerebral hemisphere and vice versa.

### Heterogeneous brain-wide associations of dyslexia disposing genetic loci

42 individual genomic loci were significantly associated with dyslexia after genome-wide multiple testing correction in the 23andMe Inc. GWAS^30^. For 35 of these loci the lead SNP in the UK Biobank data passed our quality filtering (Methods). We mapped the brain-wide associations of each of these 35 SNPs separately, with reference to increased dosage of the disposing alleles, and with no adjustment for head size for the voxel-based analysis, to avoid collider bias.

The brain-wide association maps for these 35 genetic variants showed some limited convergence, most notably in a left hemisphere medial prefrontal region peaking in Brodmann area 32^44^ that was associated with 6 of the variants, but there was also much divergence across the 35 maps (Supplementary Figure 4, Supplementary dataset). For example, for voxel-wise volumetric analysis, the locus intronic to *PPP2R3A* (index SNP rs13082684, which was the most significant dyslexia-disposing variant in the 23andME GWAS) was associated with lower volume in the right posterior insula and Heschl’s gyrus, and deep subcortical structures spanning the internal capsule, anterior thalamus, and anterior thalamic radiations (Supplementary Figure 4). The intronic *BCL11B* variant (rs35131341) was associated with lower volume in medial fronto-parietal areas, bilateral Heschl’s gyri and planum temporale (Supplementary Figure 4). In contrast, association in the opposite direction (i.e. increased volume with the predisposing allele) was observed in posterior cerebellum for the *BCL11B* variant (Supplementary Figure 4). The *SATB2* locus (rs6435017) exhibited lateralized association with lower volume of the posterior insula and left Heschl gyrus, and higher volume in the right anterior middle temporal gyrus (Supplementary Figure 4). The *AUTS2* variant (rs3735260) was associated with higher volume in the optic radiation close to the primary visual cortex, with a more pronounced effect in the right hemisphere, while the *SH2B3* locus (rs7310615) was also associated with higher volume in the right optic radiation, as well as increased volume in the left Heschl’s gyrus (Supplementary Figure 4). Brain-wide volumetric association maps for all 35 genome-wide significant dyslexia-disposing variants are in supplementary dataset.

In terms of white-matter microstructure, again the 35 genetic variants had mostly distinct brain-wide association maps (Supplementary Fig. 5). For example, a variant upstream of *NEUROD2* (rs12453682) was associated with lower apparent fiber density in tracts passing through the internal capsule and caudally extending to the brainstem and superior cerebellar peduncles (Supplementary Fig. 5). Other negative associations were observed in the superior longitudinal fasiculi for the *SH2B3* (rs7310615) and *SEMA3F* (rs2624839) loci (Supplementary Fig. 5). In contrast, positive associations were observed in the occipital lobes for the *SEMA3F* (rs2624839) and *ARFGEF2* (rs11393101) loci, such that higher apparent fiber density in the forceps major tracts were associated with the dyslexia-disposing alleles (Supplementary Fig. 5). Fixel-wise associations for all 35 genome-wide significant dyslexia-associated variants are in supplementary dataset.

### Impact modes reveal latent structure in imaging genetic associations

As described in the previous section, 35 genetic loci that were individually associated with dyslexia at a genome-wide significant level had largely distinct, but sometimes overlapping, associations with regional brain volumes or white matter microstructure. We opted to broaden this insight to thousands of genetic variants that contribute to the polygenic disposition to dyslexia, through a novel application of probabilistic independent component analysis^36^. We aimed to understand whether, despite heterogeneity of brain-wide associations for different genetic variants, there exists a latent multivariate structure. This new approach goes beyond the standard PGS approach that aggregates all disposing variants into a single scalar score per subject.

We first mapped the brain-wide associations in the UK Biobank data for each of 13,766 genetic variants that were associated with dyslexia in the 23andMe GWAS^30^ with point-wise association *P* values of less than 0.01 and clumped for linkage disequilibrium (Methods). Together, these variants contribute much of the genome-wide polygenic disposition to dyslexia. We then concatenated the resulting 13,766 brain-wide association maps and decomposed them into ten independent components, separately for voxel- and fixel-wise data (see Methods). Each component reflects a spatially independent map of brain regions associated with a distinctive set of genetic variants that exhibit similar brain-wide effects. We call these components *impact modes*, a term we partly borrowed from Smith et al. 2012^45^.

For both voxel- and fixel-wise data, impact modes localized to anatomically coherent features and were more spatially homogenous than the univariate brain maps associated with dyslexia PGS (Supplementary Fig. 6). For example, in the voxel-based volumetric data, impact mode #5 mapped distinctly to the occipital lobes and posterior thalami (Supplementary Fig. 6). Among genetic variants that contributed especially to this impact mode, a dyslexia-disposing variant at the DAAM1 locus was associated with increased volume of the primary visual cortex (rs36065072, mode weight z-score=6.2). A further example to illustrate: impact mode #8 exclusively captured bilateral associations with temporal lobes (Supplementary Fig. 6), and an intronic variant in *MAPT* was among those that exhibited a strong weight in this impact mode (rs12150530, mode weight z-score=-4.7). The weights of all 13,766 variants for the ten volumetric impact modes are provided in Supplementary Table 2.

Impact modes for fixel-based white-matter microstructure corresponded to groups of identifiable tracts. For example, diffusion impact mode #5 captured microstructural variations in the forceps major, optic radiation, and superior longitudinal fasciculus, whereas impact mode #10 isolated the internal capsule and brainstem tracts that caudally enter superior cerebellar peduncles (Fig. 3). While both of these sets of tracts were associated with the overall dyslexia PGS (Fig. 2), their association with two independent modes indicates that they stem from distinct genomic underpinnings. The genetic variant with the strongest weight for the internal capsule impact mode (#10) was upstream of *SLC39A8* (rs35518360, mode weight z-score=-7.6) (Fig. 3). This variant was only weakly associated with dyslexia (GWAS p=0.001), but has also been significantly associated with schizophrenia^46^ and educational attainment^47^. The variant is in almost full linkage disequilibrium (r^2^=0.9) with a missense variant (rs13107325) in the same *SLC39A8* gene, suggesting that an amino-acid change in the encoded protein affects microstructural properties of the internal capsule tracts. The dyslexia-disposing *NEUROD2* variant (rs12453682) exhibited the second-strongest weight for this same impact mode (#10) (mode weight z-score=5.3) (Fig. 3). The complete weights of the 13,766 genetic variants for all ten diffusion impact modes are provided in supplementary Table 3.

**Figure 3.**
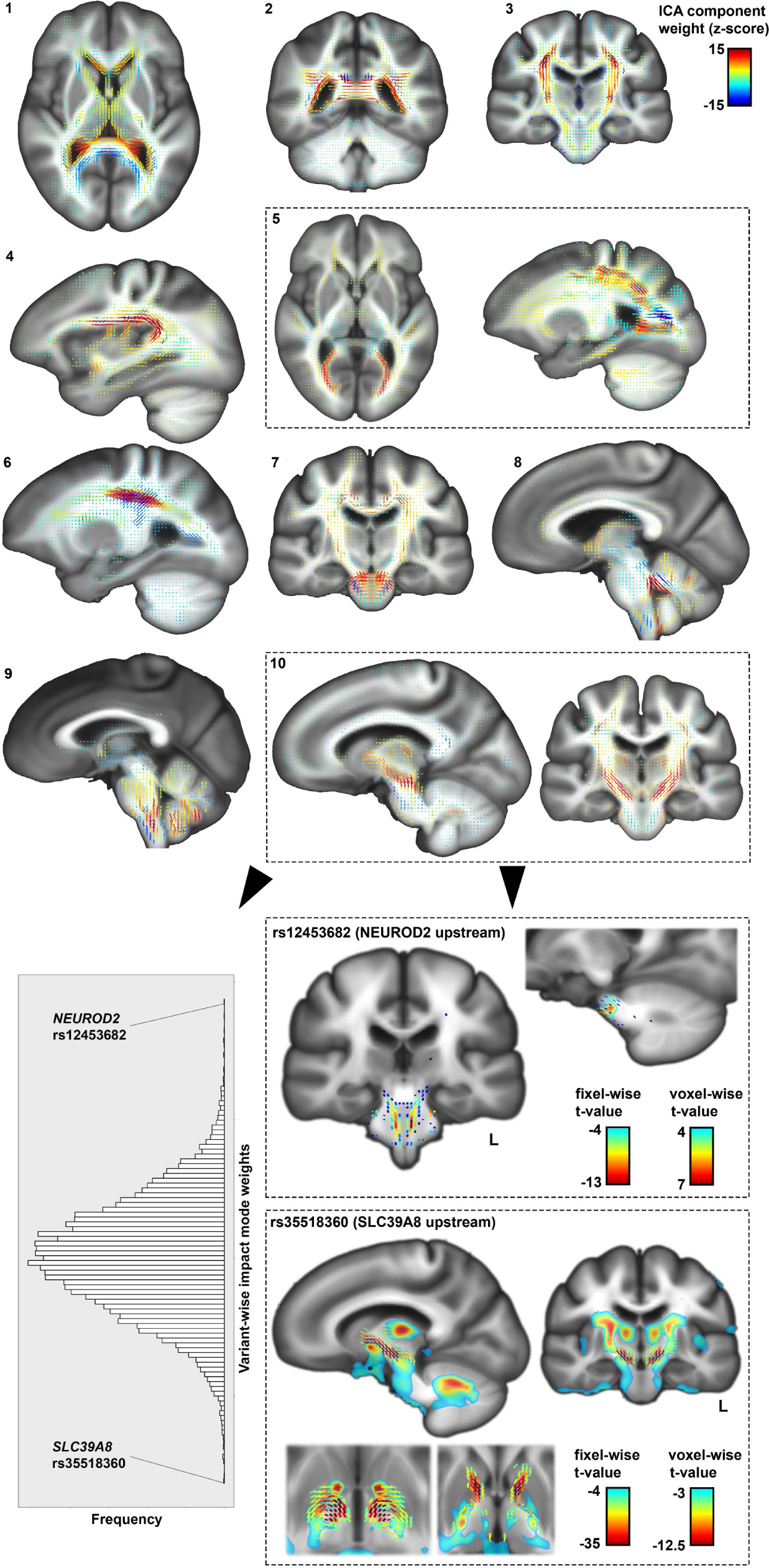
Distinct impact modes identified by independent component analysis of fixel-wise associations, for 13,766 variants that contribute to genome-wide polygenic disposition to dyslexia (top). Histogram of variant-wise weights for impact mode #10, and univariate maps of the two top loci contributing to this impact mode, *NEUROD2* and *SLC39A8*, as examples of how an impact mode can be queried in terms of specific genetic contributions (bottom).

### Overlap in brain-wide associations of dyslexia polygenic scores and genetically-correlated traits

Consistent with previous reports^30, 31^, using GWAS summary statistics from large-scale genetic studies of other cognitive, educational and behavioural traits (see Methods), we reproduced significant SNP-based genetic correlations between dyslexia and GCSE education (General Certificate of Secondary Education in the UK) (r_g_=-0.50), verbal-numerical reasoning (r_g_=-0.49), the first principal component of school grades (r_g_=-0.39), ADHD (r_g_=0.40), word reading ability (r_g_=-0.69), non-word reading ability (r_g_=-0.71), spelling (r_g_=-0.75) and phonemic awareness (r_g_=-0.62) (all with *P*-values <10^-23^) (Supplementary Fig. 7).

We then generated PGS for each of these traits in the UK Biobank imaging genetic dataset using lassosum2^40^ (the same method as for our primary analysis of dyslexia PGS above). PGS for several of these traits were associated with the volumes of basal ganglia, thalamus, and adjacent white matter tracts, especially in the internal capsule (Fig. 4). These associations extended to the frontal lobes for the fluid intelligence PGS, education PGS and school grade PGS, and more caudally to the brainstem for ADHD PGS. The directions of effects were consistent across traits, with polygenic disposition to poorer performance, lower education, and ADHD associated with lower regional volumes in these areas (Fig. 4), similarly to dyslexia PGS (Fig. 1).

**Figure 4.**
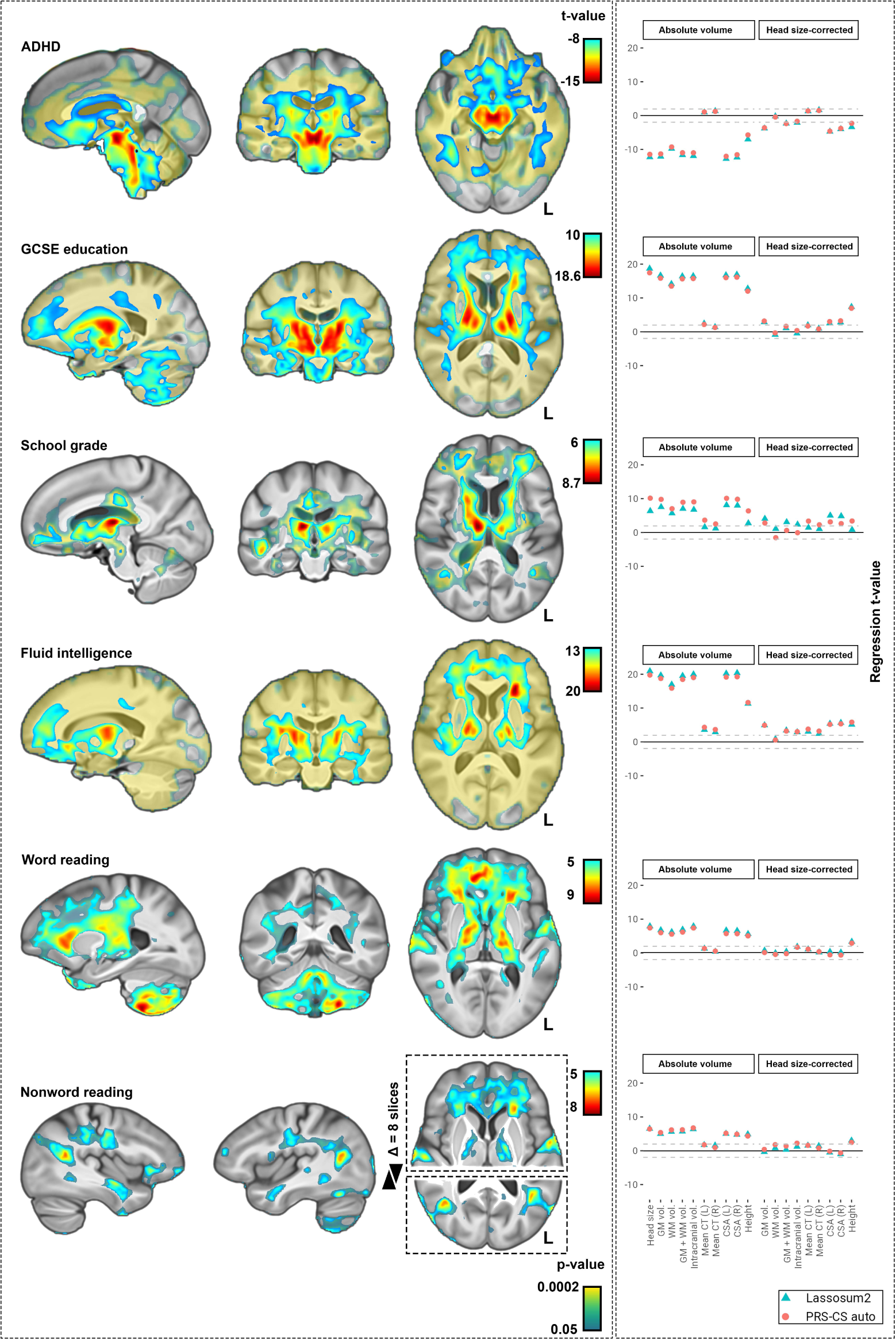
Brain-wide associations of polygenic scores (PGS) for traits that are genetically correlated with dyslexia. The panel on the left shows the associations of PGS with regional brain volumes. The panel on the right shows the associations of PGS with global imaging measures, before and after adjustment for head-size as a covariate. PGS for phonemic awareness was not significantly associated with the volume of any voxel, and therefore not shown in the figure. The PGS for spelling ability only showed a significant association in the putamen and is also not shown in this figure (its map is shown in supplementary figure 5). GM: gray-matter; WM: white-matter; CT: cortical thickness; CSA: cortical surface area. PGS: polygenic score. IDP: imaging-derived phenotype.

A lateralized association between higher dyslexia PGS and lower left anterior insula volume (Fig. 1) was also observed for PGSs for lower fluid intelligence, word reading, non-word reading, and GCSE education (Fig. 4). Lower temporoparietal junction volume, which was observed in individuals with higher dyslexia PGS (Fig. 1), was also observed in individuals with lower non-word reading PGS (Fig. 4). In contrast, as a feature that was only found for dyslexia PGS, the association with Brodmann area 4 (primary motor cortex) (Fig. 1) was not observed as a focus for PGS of other traits (Fig. 4). PGS for word reading and non-word reading were positively associated with the bilateral volumes of Heschl’s gyri that contribute to primary auditory cortex, as well as posterior cerebellum (Fig. 4).

No associations were observed between PGS for lower performance in any trait and higher regional brain volume, with the exception of spelling, for which individuals with polygenic disposition to lower performance showed higher volume in the putamen (Supplementary Fig. 8).

In white-matter fixel-based analysis, associations were observed between apparent fiber density in the internal capsule tracts and PGS for all traits (Fig. 5). Specifically, lower apparent fiber density was consistently associated here with polygenic disposition to lower performance/achievement and higher risk for ADHD. PGS for fluid intelligence and GCSE education exhibited the most extensive associations here, passing through the anterior limbs of the internal capsule (Fig. 5). In particular, the genu and anterior limb of the internal capsule emerged as a hotspot, where PGS for dyslexia and all other genetically correlated traits overlapped in their associations (Fig. 5). In addition, lower PGS for GCSE education and higher PGS for ADHD were associated with lower apparent fiber density in cerebellar dentate nuclei, similarly as for higher dyslexia PGS.

**Figure 5.**
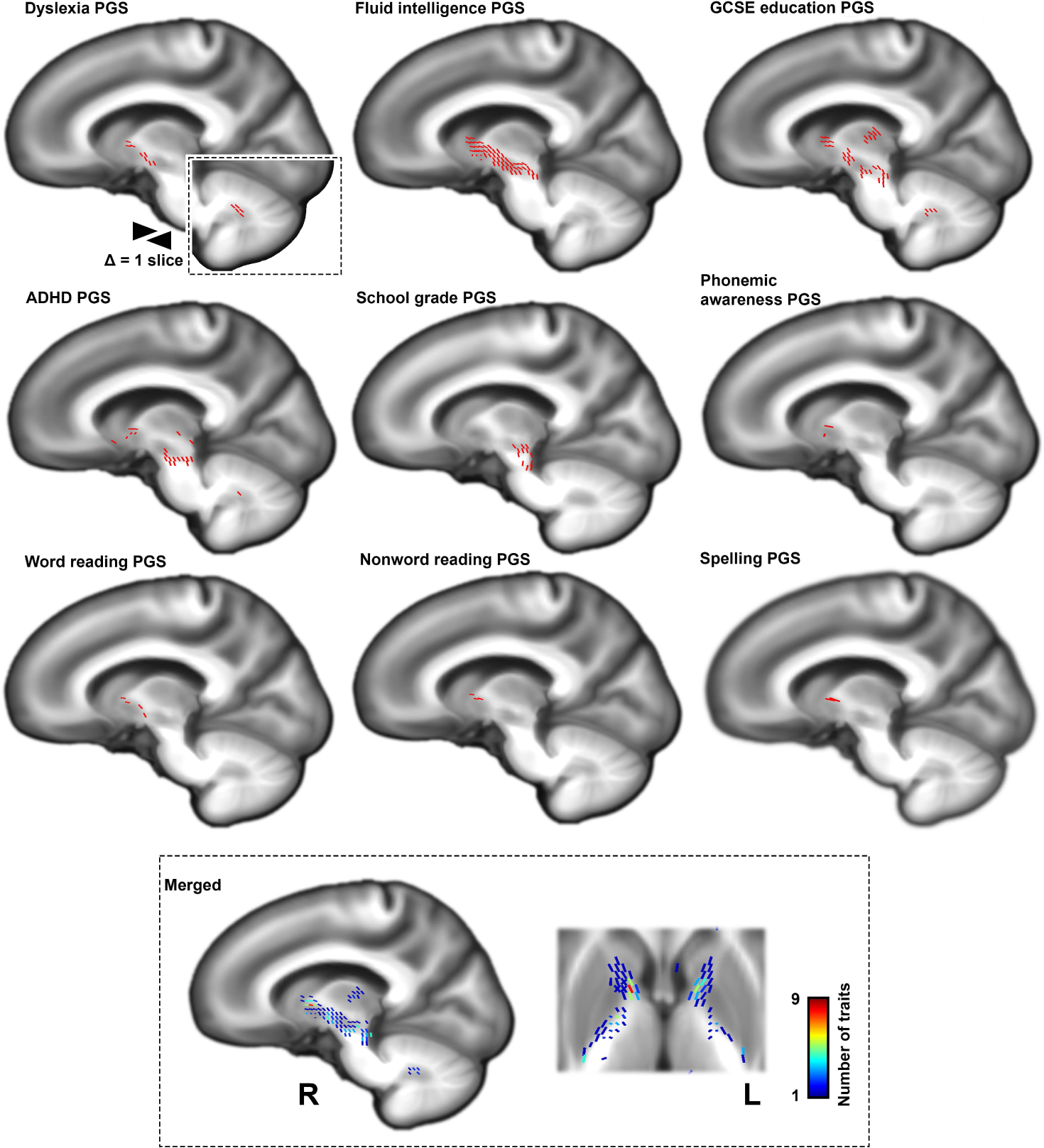
Polygenic scores (PGS) of additional traits that are genetically correlated with dyslexia show associations with white-matter microstructure, measured by apparent fiber density. Lower apparent fiber density in the internal capsule was consistently associated with polygenic disposition to lower performance/achievement and higher risk for ADHD. The genu of the internal capsule was a hotspot of shared association with PGS for all traits. PGS: polygenic score.

## Discussion

Our study of genetic disposition to dyslexia in over 30,000 adults implicated diverse brain structures, notably involved in motor, language-related and visual functions. Our sample size was more than two orders of magnitude larger than any imaging case-control study of dyslexia published to date, which is likely to have aided in robustness of our findings. Nonetheless, a direct comparison cannot be made to case-control studies. Rather, our study shows the utility of a complementary approach to studying the neurobiology of dyslexia, through identifying neural correlates of genetic disposition while leveraging large-scale population data to overcome statistical uncertainty.

A novel application of independent component analysis identified various impact modes comprising sets of dyslexia-disposing genetic loci associated with distinct brain features. This heterogeneity is consistent with dyslexia as a high-level behavioural outcome, with no simple mapping to any single brain structure, network, cognitive function or genetic factor. Dyslexia emerges from a complex interplay between genes, environmental exposures, and neural adaptations during reading acquisition^2, 11, 12, 28^, and is associated with educational and socioeconomic outcomes^48, 49^. Some of the structural brain correlates of polygenic disposition in the adult population may therefore be linked with the development of dyslexia as potential causal factors, while others might be consequences of lifestyle differences, for example time spent reading professionally or personally.

Several of the implicated brain structures were also associated with genetic dispositions to other traits including educational attainment, fluid intelligence, ADHD, and reading- and language-related performance measures across the population. However, the volume of a large continuous region along the medial wall, spanning parietal and frontal cortices and peaking within the primary motor cortex^50^, showed an association only with genetic disposition to dyslexia among all of these traits. A combination of reduced primary motor cortex volume together with alterations of other regions implicated by this study may therefore dispose individuals to dyslexia in particular. Perhaps consistent with this, children with dyslexia have shown overactivation of the primary motor cortex during reading or reading-related tasks^51^. Furthermore, at the phenotypic level, dyslexia is associated with motor difficulties^52, 53^, although many children with dyslexia show no motor impairments, and lower performance of sequential motor tasks has also been reported for ADHD^52, 53^.

Lower volume of the medial wall region spanning the primary motor cortex was notably associated with the dyslexia-disposing allele of the *BCL11B* locus. This allele was also associated with higher volume of the posterior cerebellum. Consistent with this, *BCL11B* encodes a zinc finger protein transcription factor and is expressed in cerebral cortical layer V projection neurons that send motor connections to the brainstem and cerebellum^54–57^. *BCL11B* may therefore modulate the topology of cortico-cerebellar pathways. Rare missense variants of *BCL11B* are associated with speech impairment, developmental delay and intellectual disability^58^.

Of further relevance in terms of motor circuits, a consistent finding across all of the PGSs that we studied, including dyslexia PGS, involved microstructure of the internal capsule. A clue to the role of this deep white matter tract in dyslexia is provided by diffusion impact mode #10, which isolated the internal capsule and the cerebellar dentate nuclei together as one single component, linked to a shared set of dyslexia-disposing genetic variants. Motor projections such as the dentate-thalamic tracts pass through the internal capsule, while dentate nucleus lesions are the cause of cerebellar cognitive affective syndrome (CCAS) that involves linguistic, executive and visual-spatial impairments^59^. Especially the internal capsule’s anterior limb was the main focus of convergence across the various PGS that we studied. This region reciprocally connects the thalamus and frontal cortex and is engaged in multiple cognitive domains that contribute to psychiatric traits^60^.

Specific genetic loci that contributed strongly to the internal capsule and dentate impact mode #10 included the *SLC39A8* and *NEUROD2* loci. *SLC39A8* encodes a metal ion transporter that modulates neurotransmitter receptor glycosylation^61^, and this locus has also shown genome-wide significant associations with schizophrenia^46^, intelligence^62^ and educational attainment^63^. *NEUROD2* codes for a neuronal migration and differentiation factor and its expression co-localizes with *BCL11B+* layer V pyramidal neurons^64^. In mice, *NEUROD2* knockout selectively increases excitability of layer V neurons^64^, while in humans haploinsufficiency of *NEUROD2* is associated with intellectual disability, autism and speech delay^64^. Taken together with our findings across PGS for dyslexia and various other genetically correlated traits, the internal capsule and cerebellar dentate nucleus may be involved in these traits through altered cortico-cerebellar circuits that require layer V projection neurons, with consequences for diverse aspects of cognition, including those required for normal reading.

In line with the implication of left-lateralized language-related brain regions by some previous studies of dyslexia (see the Introduction), we found that higher dyslexia PGS was associated with lower regional volume in the left temporoparietal junction and left anterior insula. The left temporoparietal junction is involved in the processing of syntactic and semantic domains of language, and coding and retrieving speech sounds^65–67^. The PGS for nonword reading and school grades were also associated with temporoparietal junction volume, further supporting the relevance of this brain region for phonological decoding ability and educational outcomes. Regarding the anterior insula, this region is closely connected to the adjacent inferior frontal gyrus^68, 69^, another core region of the language network. Insular activation recently emerged as a focus of convergence in a meta-analysis of functional MRI studies using rapid naming, rapid word reading and rapid sentence reading tasks^70^. The left anterior insula was also the peak of brain-wide association with the fluid intelligence PGS in our study. This region may therefore contribute to dyslexia in terms of both rapid reading fluency and more generally through higher cognitive processes.

We found that several of the individual genetic loci that dispose most significantly to dyslexia were associated with the volumes of primary auditory cortices (Heschl’s gyri), including *PPP2R3A, BCL11B, SATB2,* and *SH2B3*. This supports an involvement of primary auditory cortex in the neural basis of dyslexia, as has been discussed previously^71^. However, the effects that we observed were different across the genetic loci in terms of directions of effect (volumetric increases or decreases) associated with the dyslexia disposing alleles, even within the same hemisphere. This pattern might arise because altered Heschl’s gyrus volumes may be only secondary to the molecular and cellular roles of these genes in auditory cortex function that are relevant for dyslexia.

Regarding visual circuits, increased polygenic disposition to dyslexia was associated with increased apparent fiber density in the forceps major white matter tract. Lesions of the forceps major, which connect bilateral occipital cortices, lead to topographical disorientation in humans^72^. In addition, the *AUTS2* dyslexia-disposing variant was associated with higher volume in the optic radiation. Our findings may relate causally to visuo-orthographic deficits in dyslexia^73^, or might signify a secondary adaptation of the visual network to lower reading activity in adults with higher genetic disposition (for example due to reading avoidance). We also saw a non-significant trend towards increased volume of primary visual cortex in those with higher polygenic disposition to dyslexia, which became significant after adjusting for head size in a secondary analysis. As noted in the Results, adjusting for head or brain size risks introducing collider bias whenever polygenic disposition and regional volume both influence total brain volume, which seems a likely scenario. We therefore regard the head size-adjusted results with caution and advocate more awareness of collider bias in brain imaging genetic studies, especiallywhen including covariates that are themselves heritable.

Our study has several limitations. Although the UK Biobank is a population sample, this cohort is healthier on average than the general UK population due to volunteering bias^74^. Dyslexia PGS was derived using data from a large GWAS study of participants who self-reported having received a diagnosis of dyslexia^30^, but with no information on the type, timing or severity of this condition (e.g. no distinction was made between acquired and developmental dyslexia, and no information was recorded on who had made the diagnosis). Nevertheless, strongly negative genetic correlations of this dyslexia phenotype with quantitative measures of word and non-word reading, spelling and phoneme awareness argue for its validity^30, 31^. The present cross-sectional study was carried out using adult data, which means that cause-effect aspects are not possible to disentangle with certainty. Future large-scale imaging genetic studies would benefit from longitudinal data from children, to inform on genomic impact modes for structural brain changes during reading acquisition.

In summary, we identified multiple brain networks linked to genetic disposition to dyslexia in a large adult sample from the general population. This approach complements classical case-control designs, for which it has not been possible to collect sample sizes within the same order of magnitude as that used here. Our study revealed that genetic disposition to dyslexia can be broken down into distinct sets of factors that associate with various identifiable brain networks, consistent with dyslexia as a complex and heterogeneous trait. Our study also showed which brain structural features are associated in common across multiple traits that are genetically correlated with dyslexia, as opposed to being associated with dyslexia genetic disposition alone among these traits.

### Methods Genetics

UK Biobank data were accessed following approval of the application number 16066, P.I. Clyde Francks. UK Biobank is an in-depth investigation of more than 500,000 volunteers in the UK who are assessed for health, lifestyle, genomic, and many other variables^32^. Multimodal brain MRI data had also been released for approximately 10% of the individuals when the present study was initiated in 2022^34, 75^. The UK Biobank received ethical approval from the National Research Ethics Service Committee North West-Haydock (reference 11/NW/0382), and all of their procedures were performed in accordance with the World Medical Association guidelines. Written informed consent was provided by all of the enrolled participants.

Genotyping has been performed using either BiLEVE Axiom or Axiom arrays from Affymetrix, which target highly overlapping sets of ∼800,000 genomic variants with more than 95% similarity^76^. The UK Biobank has also released common genome-wide variants imputed to Haplotype Reference Consortium and UK10K haplotypes^76^. In this study we focused on participants who also underwent brain MRI at one of the four imaging sites and for which at least one usable T1-weighted and/or diffusion MRI (dMRI) scan had been produced (see the next section). The genetic analyses were focused to the largest ancestry group within this cohort, recorded as ‘white British’ using a combination of self-report and genomic principal component analysis (this group constitutes ∼85% of the overall dataset: data field #22006). Pairs of genetically related subjects with kinship coefficients above 0.044 were identified in the target sample (70). Individuals related to the largest number of others were recursively removed until no two individuals were related at or above this kinship threshold, leaving 35,231 individuals (18,363 females). The resulting sample encompassed individuals aged from 45 to 82 years, with a mean age of 64.2 years and a standard deviation of 7.7 years. We then included bi-allelic genetic variants with minor allele frequency >= 0.01, imputation quality score of higher than 0.7, and Hardy-Weinberg equilibrium p-value of greater than 10^-7^, yielding 8,366,177 autosomal single nucleotide variants (SNVs) and 1,092,696 short insertion-deletions (indels).

### Structural MRI: tensor-based morphometry

We accessed minimally processed and brain-extracted T1-weighted brain MRI volumes of 42,798 individuals^34, 75^ for tensor-based morphometry using symmetric image normalization (SyN) registration^37, 38^. For the present study we generated a study-specific average brain template in a randomly chosen subset of 1000 individuals. The template was generated through 11 consecutive Advanced Normalization Tools (ANTs v2.3.5) registrations that iteratively refined the template shape using rigid, affine, and diffeomorphic SyN transformations at incremental resolutions up to native resolution (i.e. 1mm^3^). Thereafter, all individuals’ original T1-weighted brain volumes were histogram matched, winsorized at 1-99 percentiles, and non-linearly registered to our study-specific template using SyN. Registration parameters included a variance for total field of three, and variance for update field of zero, a downsampling scheme of 6×, 4×, 2×, and 1× (i.e. full resolution) and smoothing sigmas of 4, 2, 1 and zero voxels. A cross-correlation metric with a radius of four voxels was used.

The affine registration matrix was composed with the SyN deformation field and the final warps were subsequently converted to Jacobian determinant maps, which encode the amount of regional brain tissue ‘shrinkage’ or ‘expansion’ in the brain of each individual as compared to our study-specific, average template. ANTs affine registrations failed in 2,098 individuals; instead of removing them, we opted to use a comparable linear registration method, FSL Flirt^77^ to initialize SyN, while controlling for a potential batch effect in subsequent analyses as a binary covariate.

### Diffusion MRI: data preprocessing and fixel-based analysis

We retrieved minimally-preprocessed dMRI volumes of 37,930 subjects from UK Biobank^34, 75^. These data have been collected at 2mm^3^ isotropic resolution across 100 different diffusion-encoding directions evenly distributed on two spherical shells at b-values of 1000 and 2000 s/mm^2^, as well as eight blip-reversed b≅0 volumes. Diffusion images have been corrected for off-resonance warps, gradient non-linearity, Eddy currents, and head motion by the UK Biobank team^34, 75^. For the present study we reran these corrections on raw data for a first batch of 8,247 individuals whose corrected b-vector tables were not available, while accounting for a potential batch effect in the subsequent regression model fits through use of a binary covariate. After data preprocessing, we constructed a study-specific fiber orientation density (FOD) template using MRTrix3 v3.0.3^78^ from a random subset of 890 individuals who passed registration quality control by visual inspection out of 1000. This procedure started by N4 bias field correction and intensity normalisation of the preprocessed diffusion volumes, and estimation of the average tract response function^79^. Thereafter, spherical deconvolution was performed using the estimated response function to generate subject-wide FOD volumes. These volumes were subsequently non-linearly registered to a common space and an average FOD template was generated iteratively. The FOD template was then ‘fixelated’ to identify the principal directions of white-matter tracts in each voxel. The same procedures were repeated in all 37,930 individuals to generate FOD volumes, which were then registered to the study-specific FOD template^78^. FOD registrations passed quality control in 37,884 individuals following visual inspection of each individual’s template-transformed zeroth-order harmonic map, representing average isotropic diffusion in each voxel. FOD volumes were segmented to obtain fixel-wise readouts, which were then transformed to the template’s fixel-wise space^78^. We considered apparent fiber density (AFD) readouts as a measure of white-matter microstructure for subsequent analyses^39^. In combination with genetic data, the sample available was 31,695 adult individuals (16,198 female).

### Optimizing the polygenic scoring for imaging genetic analysis

We first concatenated the voxel-wise Jacobian and fixel-wise AFD maps across all individuals and then applied MELODIC independent component analysis^80^ to extract imaging derived phenotypes (IDPs). MELODIC was performed separately per each imaging modality and at various dimensions to extract IDPs at incremental levels of spatial detail, following a geometric series corresponding with dimensions 11, 18, 29, 47, 76, 124, 200 and 324. Due to the large size of this data matrix (6.2×10^10^ voxels in structural MRI, one terabyte), we used 8,000 internal eigenmaps for independent source decomposition^81^. In addition, principal component analysis was performed on the same data and the first 324 principal components were extracted as additional IDPs. Altogether, a total of 1,153 IDPs were extracted from voxel-wise Jacobian maps and an equal number of IDPs from the fixel-wise AFD data. These IDPs were derived for the purpose of optimizing our polygenic scoring, but they were not used for our voxel-or fixel-based imaging genetic analyses, nor our impact mode analysis, which form the bulk of the findings in this study.

We used summary statistics from the largest genome-wide association study (GWAS) of dyslexia that has been performed to date, carried out by 23andMe, Inc.^30^. This GWAS study was based on 51,800 individuals of European ancestry who answered ‘Yes’ to the question ‘Have you been diagnosed with dyslexia?’, and 1,087,070 control individuals who answered ‘No’. The SNP-wise effect sizes from this GWAS were then applied to the genotype data of UK Biobank individuals, to estimate the polygenic disposition of each UK Biobank individual to dyslexia based on the combined effects of their autosome-wide genetic variants.

Our primary approach for polygenic scoring was based on the Lassosum2 model^40^. We observed strong correlation between Lassosum2 PGS and two automated PGS methods, SBayesR^41^ and PRS-CS_auto42_. Lassosum2 explained slightly more proportion of variance in brain IDPs across PGS of all studied traits (Supplementary Fig. 1) and was therefore used to present the main results. This method fits a sparse elastic-net regression and optimizes two shrinkage penalties, including L1-norm (λ) and L2-norm (δ). A grid search across 30 λ and 10 δ values was utilized for optimization with respect to maximizing the top association with any IDP. The associations of dyslexia PGS were quantified with all 1,153 IDPs in each imaging modality using linear regression. A set of confound covariates were controlled for, including subject age at imaging visit (data field #21003, instance 2), age^2^, sex (data field #31), age×sex, age^2^×sex, the first ten principal components of genomic ancestry (data field #22009), genotyping array (data field #22000, either BiLEVE or Axiom), three dummy covariates encoding four UK Biobank neuroimaging sites (data field #54, instance 2), and the number of days passed since MRI scan incepted at the site (as a measure of slow drifts in MRI hardware performance; data field #53, instance 2). For structural MRI data, the type of affine registration (i.e. ANTs or Flirt) was further controlled as an extra covariate. Structural MRI analysis was performed with and without correction for head size scaling factor (data field #25000). For diffusion MRI data, the batch effect associated with diffusion preprocessing (i.e. either performed by our team or by the UK Biobank) was added to the covariates. We found that high δ values in the range of 10^2^-10^4^ slightly increased the accuracy of Lassosum2 over automated models PRS-CS_auto_ and SBayesR, and λ in the range of 10^-5^-10^-2^ resulted in the highest accuracy of trait prediction (Supplementary Figure 1). These shrinkage parameters were therefore used for subsequent analyses.

### Voxel- and fixel-wise brain associations with dyslexia polygenic scores

We tested the brain-wide associations of dyslexia PGS with the voxel-wise and fixel-wise data in the UK Biobank. Both parametric (fsl_glm 6.0.3^82^) and non-parametric (randomise v2.9^83^) linear regression models were fitted to the data, the former to yield t-value maps for visualization and impact mode analysis, and the latter to generate brain-wide multiple comparisons-corrected statistical maps. To reduce computation costs, voxel-wise permutations were performed at half (2mm^3^ isotropic) resolution with a wall-time of 9 days for 5,000 permutations per statistical contrast. The Randomise C++ code was modified to prevent short integer overflows due to the study sample size. No cluster enhancement was applied. The same sets of covariates as the previous section were used as for optimization. In all cases, we observed that a parametric t-score of > 4.6 was equivalent to a non-parametric brain-wide corrected p-value of smaller than 0.05.

As a check on the validity of our findings obtained with Lassosum2, we applied other methods for deriving PGS: SBayesR, PRS-CS, and PRS-CS_auto_. PRS-CS applies continuous shrinkage on variant-wise weights using Bayesian priors and is optimized using a single global shrinkage hyperparameter (ϕ). We explored four different ϕ values for optimizing PRS-CS, which were 10^-6^, 10^-4^, 0.01, and 1 (Supplementary Fig. 1). PRS-CS_auto_ and SBayesR are automated polygenic scoring methods and did not require hyperparameter optimization on an independent dataset. We found that dyslexia lassosum2 PGS was strongly correlated with dyslexia PGS derived from PRS-CS_auto_ (r=0.87 and 0.93 following optimization on structural or diffusion-derived measures, respectively) and SBayesR (r=0.74 and 0.84, same order). Compared to lassosum2, these additional PGS exhibited highly similar brain-wide associations (Supplementary Fig. 2).

To describe the white matter tracts that run through regions where fixels showed significant associations of AFD with dyslexia PGS, we ran probabilistic fiber tractography using the second-order Integration over Fiber Orientation Distributions (iFOD2) algorithm in the template space^84^.

### Dyslexia locus-based neuroimaging association

42 individual genomic loci were significantly associated with dyslexia after genome-wide multiple testing correction in the 23andMe Inc. GWAS for dyslexia^30^. 35 of these variants passed our genetic quality control process in the UK Biobank data (see the Methods section ‘Genetics’, above). At each of these 35 loci, dosage of the dyslexia disposing allele was calculated and used in separate linear regression models to find brain-wide associations with regional volume and white-matter microstructure (i.e. voxel-wise Jacobian values and fixel-wise AFD values, respectively), using the same approach and covariates as when testing voxel-wise and fixel-wise PGS associations. These covariates included age, age^2^, sex, age×sex, age^2^×sex, ten principal components of genomic ancestry, genotyping array, UK Biobank imaging site, the number of days passed since MRI scan incepted at the site, the type of affine registration (for structural MRI), and preprocessing being either performed by our team or by the UK Biobank team (for diffusion MRI).

### Impact modes

PGS approximate polygenic influences through a single scalar value. These models represent a weighted average of all disposing allele counts and are agnostic to variability in the brain-wide associations of variants. Such aggregation of risk variants may obscure heterogeneous or opposing effects across the genome. We aimed to model the heterogeneity and the hidden covariance patterns in the brain-wide genomic associations. To achieve this, we initially created a brain-wide univariate association map (i.e. voxel-wise or fixel-wise t-score maps generated by a parametric regression) for each of the top independent 13,766 dyslexia GWAS loci, after clumping at a GWAS p-value threshold of less than 0.01, linkage disequilibrium r^2^ threshold of less than 0.1 and genomic window size of 500 kb (and using the same set of covariates as in all sections above). These voxel-or fixel-wise t-score maps were then concatenated across all 13,766 variants and decomposed by MELODIC into ten independent components, separately per each imaging modality. The default MELODIC data transformations, including variance normalization and mean signal removal, were not applied as these momentums reflect meaningful signals in t-score maps^85^. The extracted independent components, henceforth referred to as genomic *impact modes*, capture the hidden sources that shape the brain-wide influence of dyslexia-disposing variants and approximate their heterogeneous spatial profiles through a limited number of features.

### Additional traits related to dyslexia

We first used LD score regression^86, 87^ to confirm that we could detect previously reported genetic correlations between dyslexia and each of eight other behavioural, cognitive or education-related traits, based on summary statistics from the 23andMe dyslexia GWAS^30^ and other large-scale GWAS studies: Attention deficit/hyperactivity disorder (ADHD^88^), verbal numerical reasoning (a.k.a. *fluid intelligence*) (Pan-UKB team. https://pan.ukbb.broadinstitute.org. 2020.), the first principal component of school grades in mathematics and language^89^, General Certificate of Secondary Education (GCSE) education (Pan-UKB team. https://pan.ukbb.broadinstitute.org. 2020.), word reading, non-word reading, spelling, and phonemic awareness^31^. All of these traits showed significant genetic correlations r_g_ > 0.4 with dyslexia in our analysis (all P < 10^-23^, supplementary Fig. 7.)

In order to compare and contrast with dyslexia PGS, we then used Lassosum2 to generate PGS in the UK Biobank data for each of these eight additional traits, and mapped their brain-wide associations with the voxel-wise and fixel-wise data, using the same approach as for the dyslexia PGS.

## Acknowledgements

This research was funded by the Max Planck Society (Germany) and the Netherlands Organization for Scientific Research (Language in Interaction consortium: Gravitation grant number 024.001.006). The Wellcome Centre for Integrative Neuroimaging (RM) is supported by core funding from the Wellcome Trust [201319/Z/16/Z]. The funders had no role in study design, data collection and analysis, the decision to publish, or preparation of the manuscript. The study was conducted using the UK Biobank resource under application no. 16066 with C.F. as the principal applicant. Our study made use of quality-controlled brain images generated by an image-processing pipeline developed and run on behalf of the UK Biobank. The authors would like to thank the research participants and employees of 23andMe, Inc. for making this work possible.

## Data availability

Applications for UK Biobank data should be made via their website (https://www.ukbiobank.ac.uk) and are available following review and material transfer agreement. The full 23andMe dyslexia GWAS summary statistics can be requested from 23andMe (https://research.23andme.com/dataset-access) and are available following review and data transfer agreement. Open-source software used in this study include FSL (https://fsl.fmrib.ox.ac.uk), ANTs toolkit (https://stnava.github.io/ANTs), MRtrix3 (https://www.mrtrix.org/), Bigsnpr (https://privefl.github.io/bigsnpr), GTCB (https://cnsgenomics.com/software/gctb), qctool (https://www.well.ox.ac.uk/~gav/qctool_v2), PRS-CS (https://github.com/getian107/PRScs), and LD score regression (https://github.com/bulik/ldsc). Data generated in this project have been provided as supplementary material and/or uploaded in NeuroVault or to the UK Biobank.

## Author contributions

Conceptualization: S.S., R.M., S.E.F., C.F.; Methodology: S.S., R.M., S.E.F., C.F.; Software: S.S.; Formal analysis: S.S.; Data curation: D.S., S.S.; Writing; original draft: S.S., C.F.; Writing - review & editing: S.S., D.S., R.M., S.E.F., C.F.; Visualization: S.S.; Project administration: C.F.; Resources: R.M., S.E.F., C.F.; Funding acquisition: R.M., S.E.F., C.F.; Supervision: R.M., S.E.F., C.F.

## Supplementary material

**Supplementary Figure 1.**
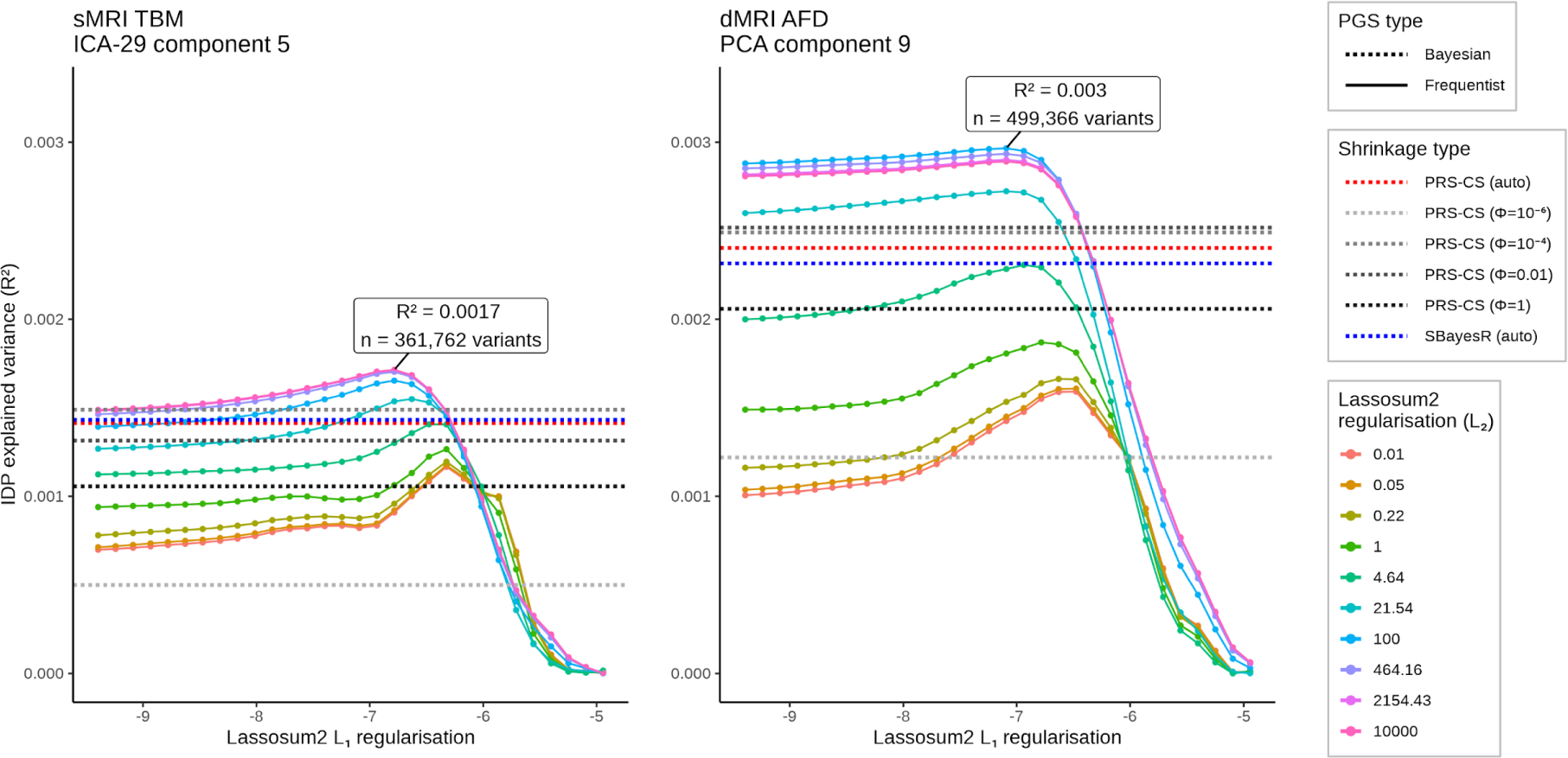
Optimization process for dyslexia polygenic score models using phenotypes derived from tensor-based morphometry (left) and fixel-based analysis (right). The Lassosum2 polygenic scores were optimized by adjusting the L1 and L2 regularization penalties. Additionally, we compare the results of two automated polygenic scoring methods, SBayesR and PRS-CS auto, represented as dashed blue and red lines, respectively. Furthermore, we explore the manual optimization of PRS-CS scores (dashed grey lines), employing four different shrinkage parameters. IDP: imaging-derived phenotype. AFD: apparent fiber density. TBM: tensor-based morphometry. PGS: polygenic score.

**Supplementary Figure 2.**
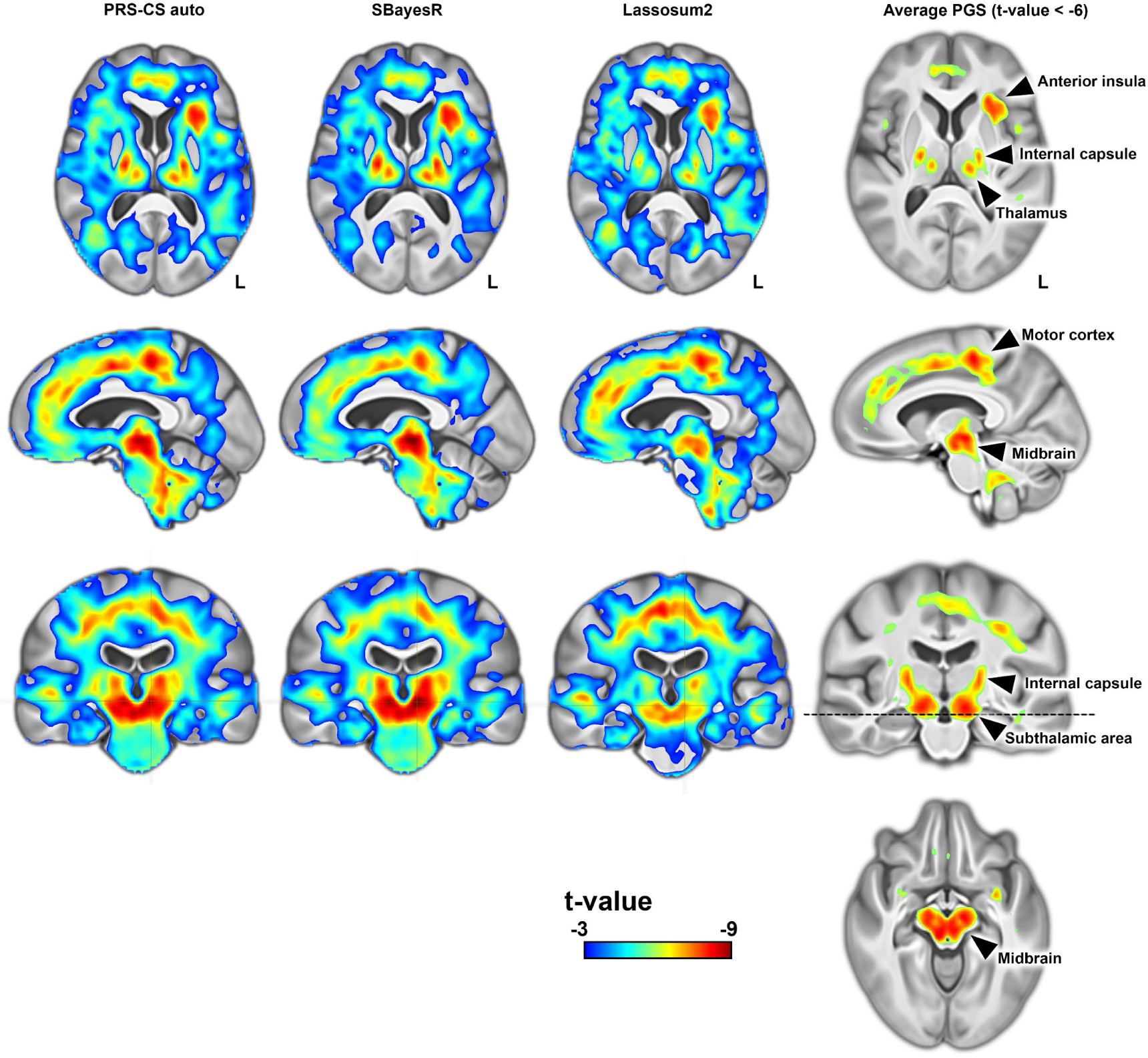
Dyslexia polygenic scores generated by three different polygenic methods (top) and their voxel-wise associations with regional brain volume (bottom) yield similar statistical brain maps. Triangles represent the coordinates of statistical peaks in an average statistical map of all three polygenic models.

**Supplementary Figure 3.**
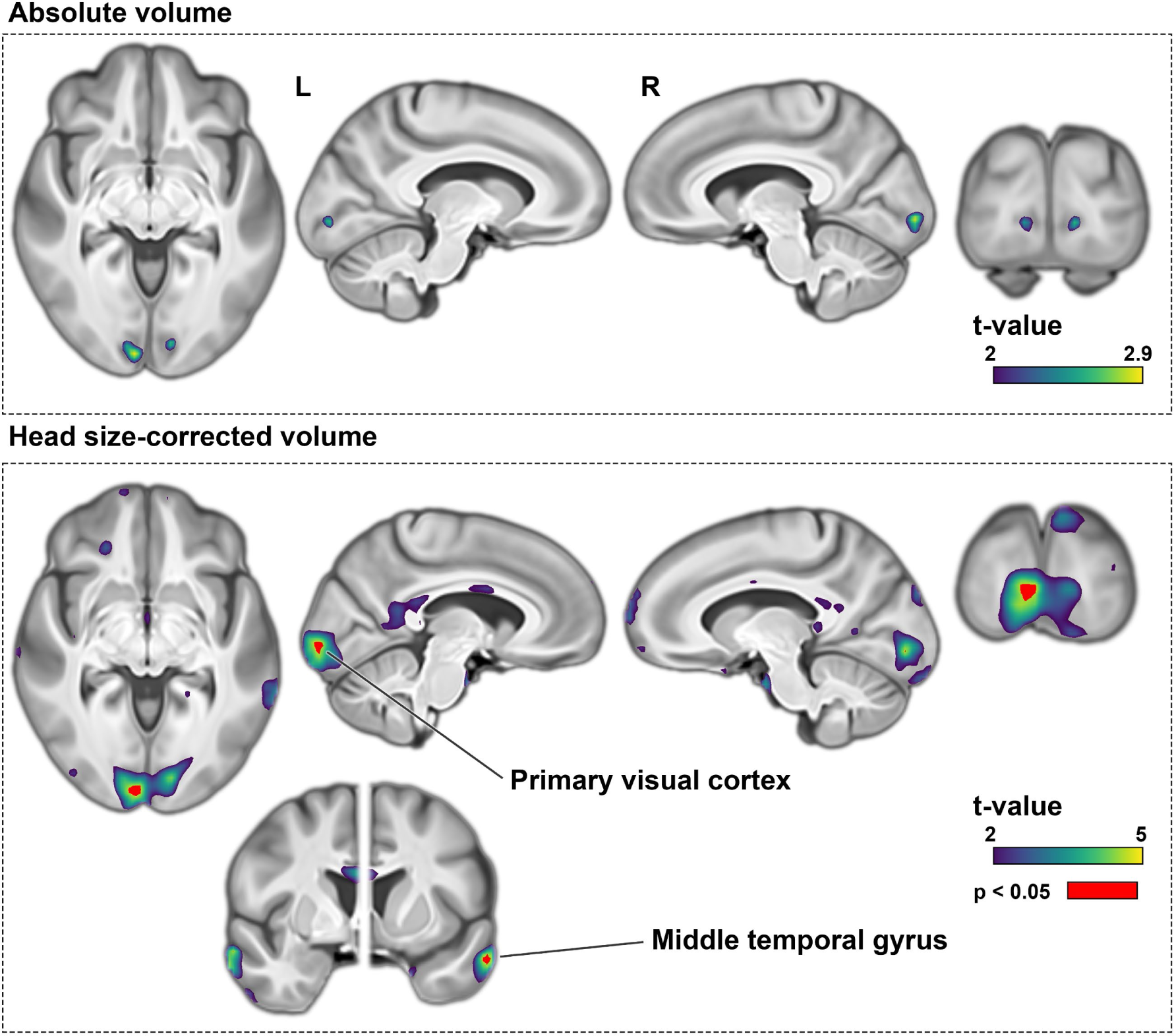
Top: Positive dyslexia PGS associations with voxel-wise volume measures before head-size correction. No voxel passed brain-wide multiple comparisons correction following 5,000 permutations before head-size adjustment. Bottom: After head-size adjustment, voxels in bilateral primary visual cortices and middle temporal gyri passed multiple comparisons significance threshold. The t-values indicate parametric regression statistics.

**Supplementary Figure 4.**
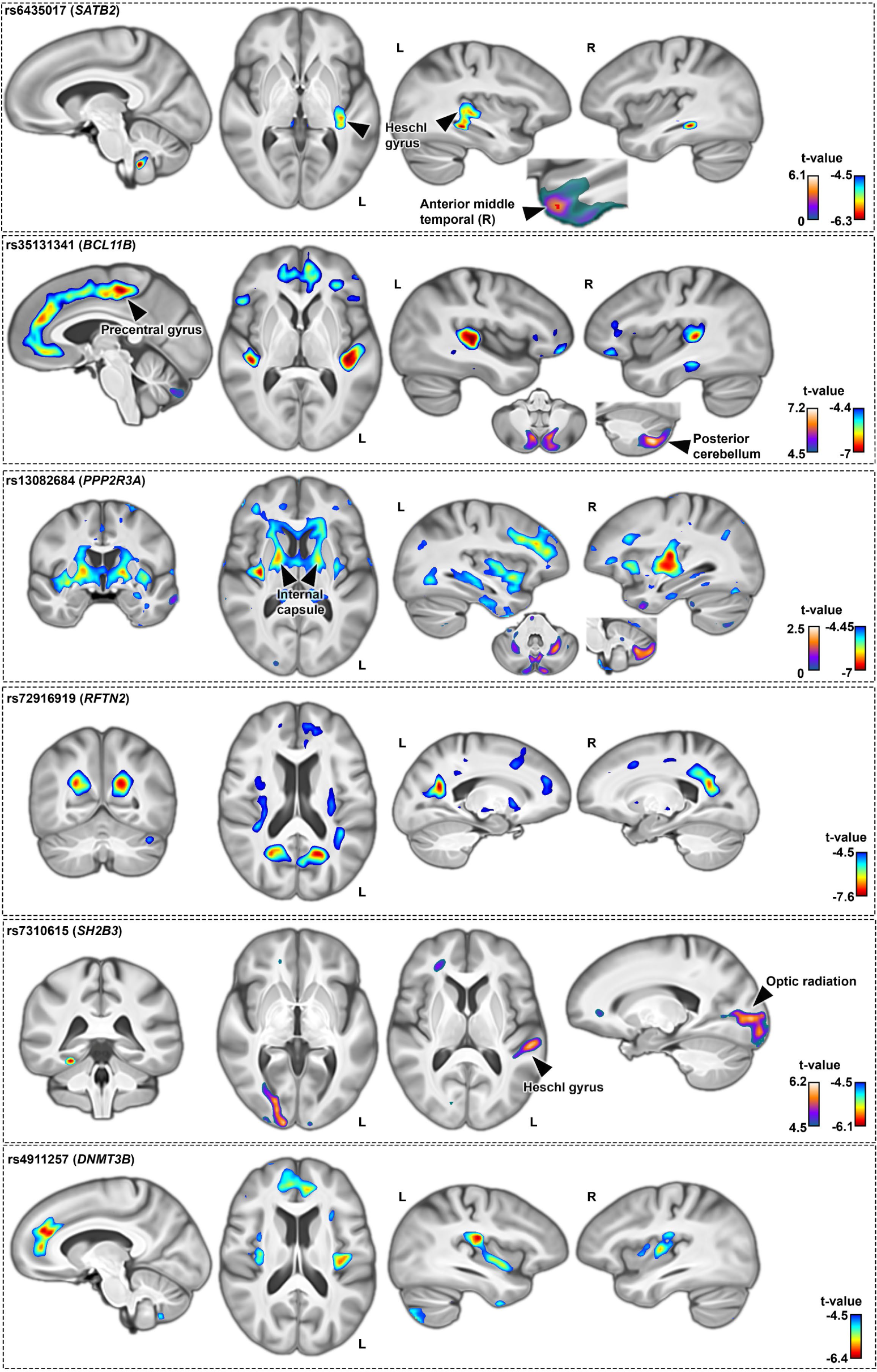
Voxel-wise associations with regional brain volume for six variants that were associated with dyslexia at a genome-wide significant level and exhibited volumetric associations surpassing 5 cm^3^. Brain maps of all 35 genome-wide significant variants are also provided in supplementary dataset.

**Supplementary Figure 5.**
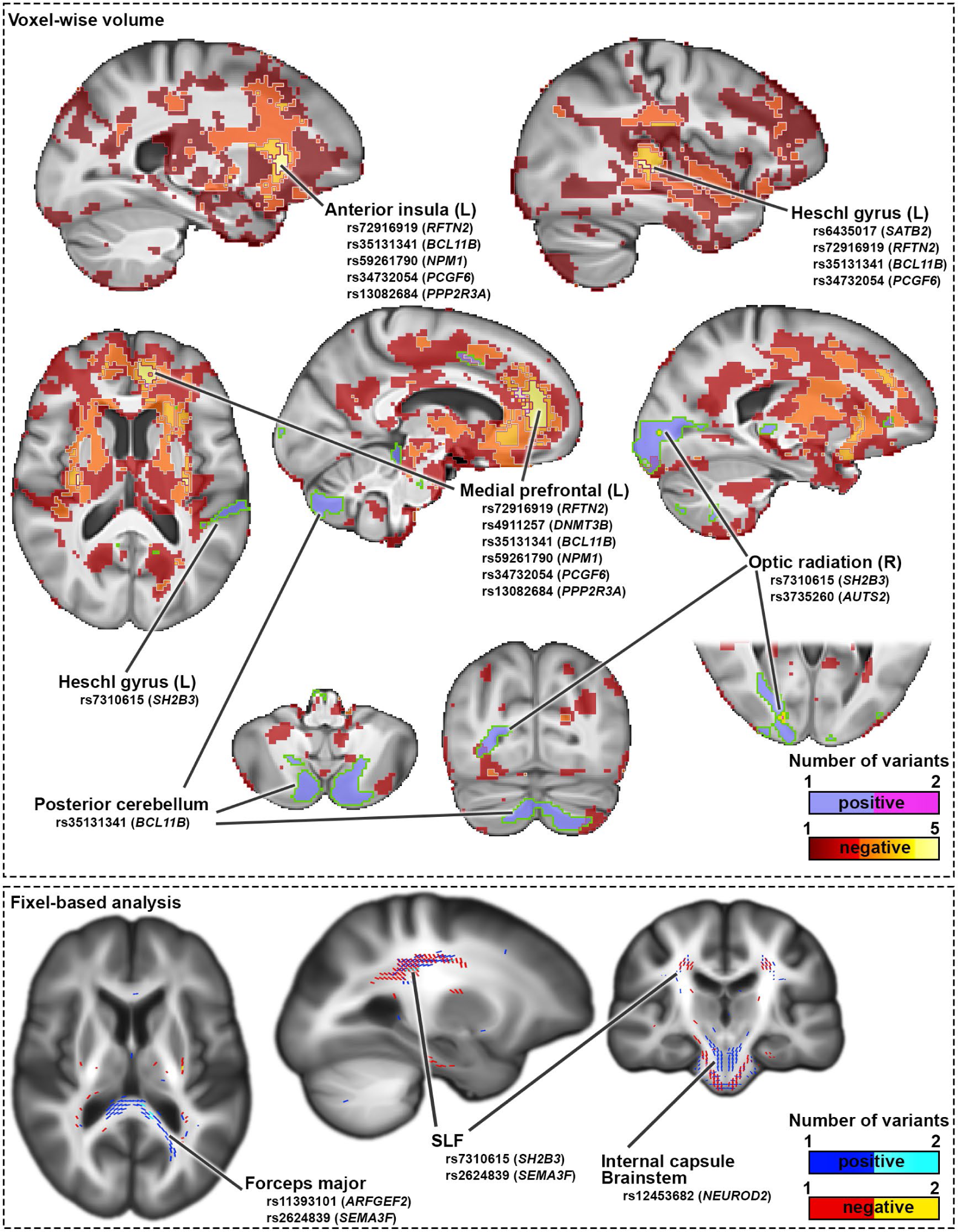
Brain-wise volumetric (top) and fixel-wise apparent fiber density (bottom) association maps in the UK Biobank data, for 35 genetic variants that were significantly associated with dyslexia at a genome-wide significant level in the 23andMe GWAS. Values indicate the number of variants at each voxel that show significant positive or negative association with respect to the dyslexia-disposing allele, as obtained from a non-parametric test following 5000 permutations. Note that the overlap in terms of affected brain regions is generally low among these dyslexia-associated variants, as the few regions of overlap involve no more than 6 of the 35 variants. Separate maps for all 35 variants are provided in supplementary dataset.

**Supplementary Figure 6.**
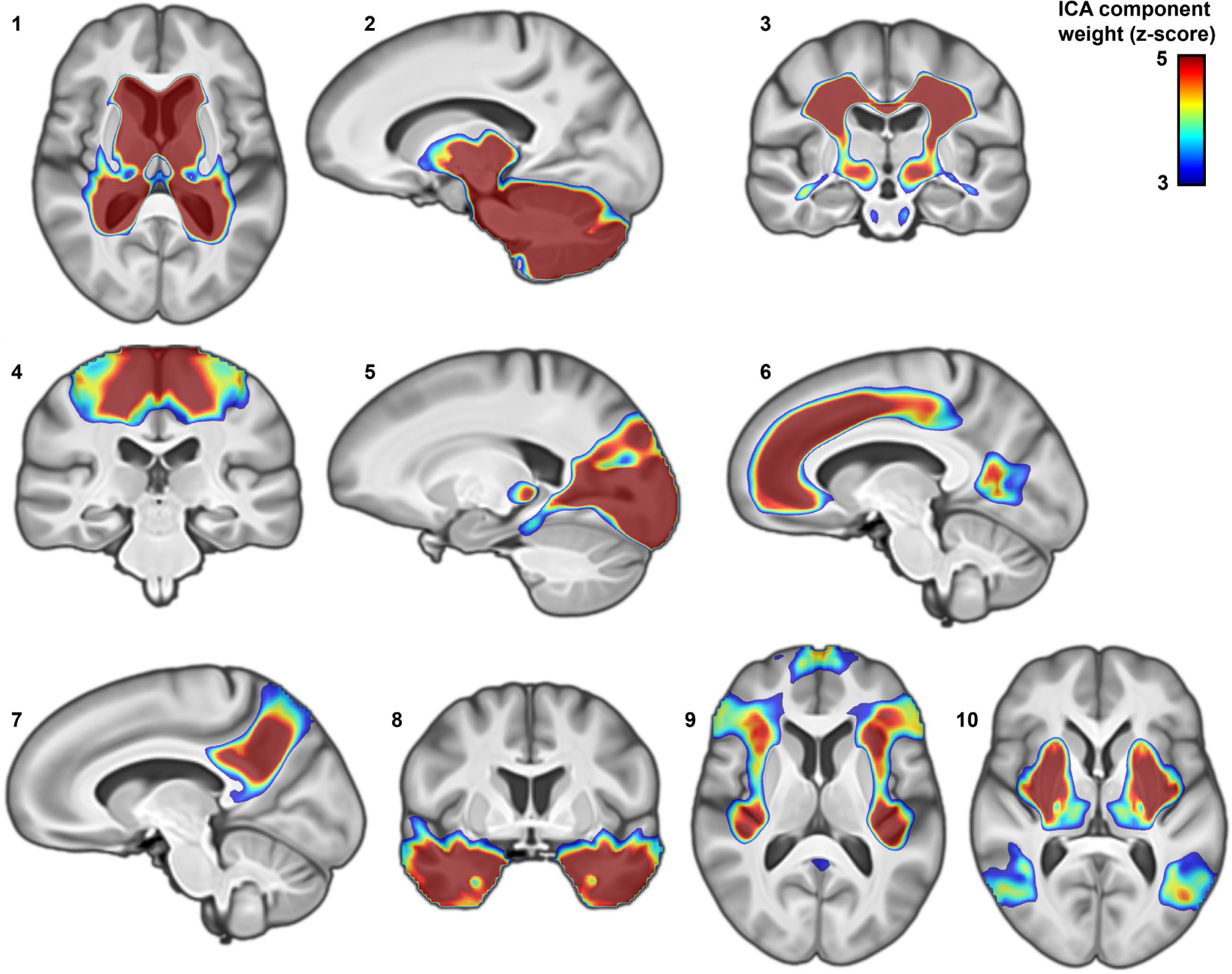
Ten genomic impact modes identified by independent component analysis of brain morphometry z-maps corresponding to the top 13,766 independent dyslexia-disposing variants. Through this analysis, the overall polygenic disposition to dyslexia can be decomposed into distinct spatial components in terms of contributing genetic variants and their specific brain-wide associations. Z-scores indicate the contribution and ‘weight’ of each voxel in the corresponding independent component.

**Supplementary Figure 7.**
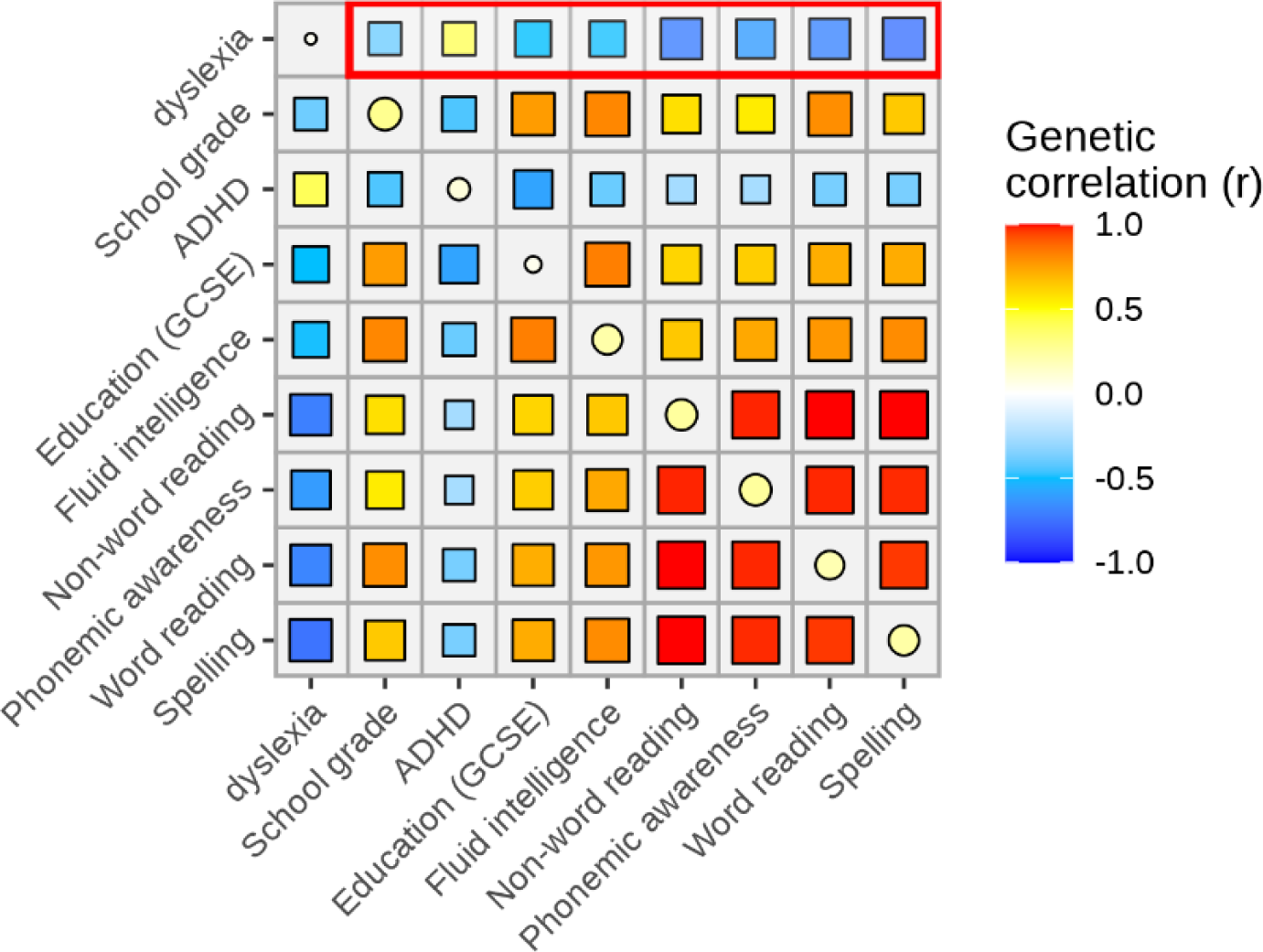
Genetic correlations of dyslexia with other traits (see Methods for the data sources).

**Supplementary Figure 8.**
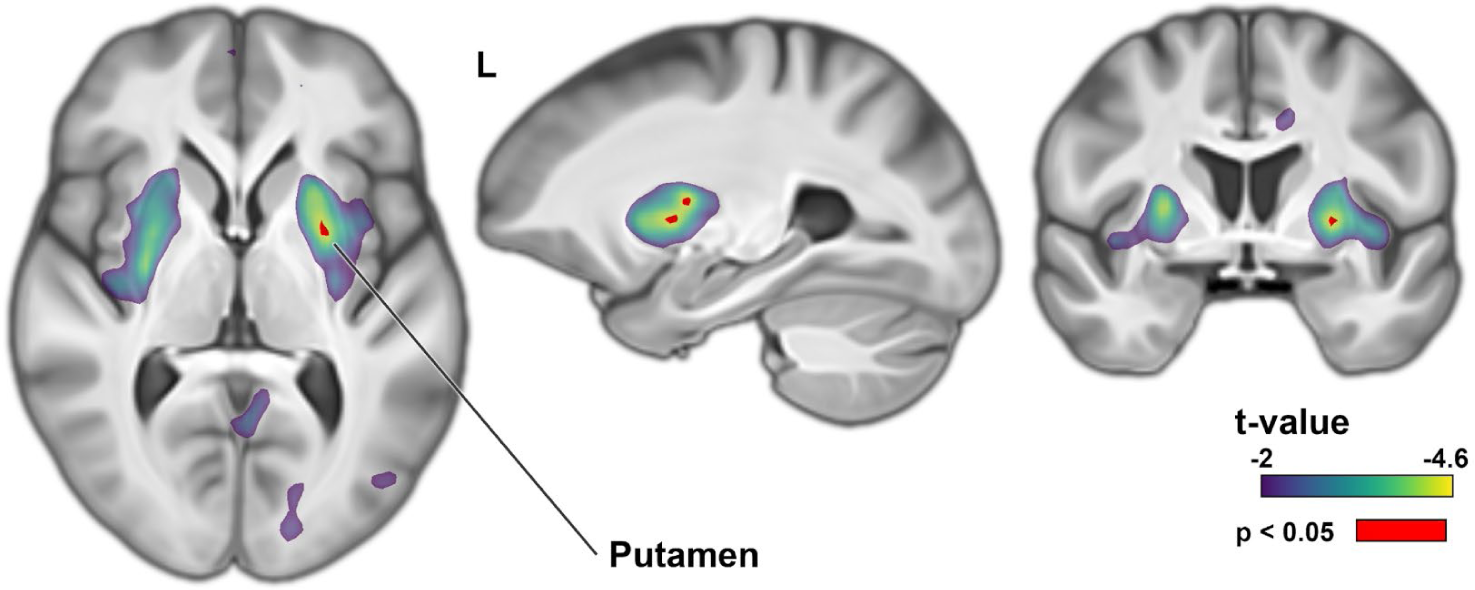
Brain-wide association of spelling performance polygenic scores. Higher PGS for better spelling performance is associated with lower putamen volume. t-values indicate parametric regression tests. Voxels passing brain-wide multiple comparisons correction following 5,000 permutations are shown in red.

